# Specific gait changes in prodromal hereditary spastic paraplegia type 4 - preSPG4 study

**DOI:** 10.1101/2022.04.20.22274071

**Authors:** Christian Laßmann, Winfried Ilg, Marc Schneider, Maximilian Völker, Daniel F. B. Haeufle, Rebecca Schüle, Martin Giese, Ludger Schöls, Tim W. Rattay

**Author notes:** Corresponding author Correspondence to: Winfried Ilg, Section Computational Sensomotorics, Hertie Institute for Clinical Brain Research Otfried-Müller-Str. 25, 72076 Tübingen, Germany.

## Abstract

**Background:** In hereditary spastic paraplegia type 4 (SPG4), subclinical gait changes might occur years before patients realize gait disturbances. The prodromal phase of neurodegenerative disease is of particular interest to halt disease progression by future interventions before impairment has manifested.

**Objectives:** Identification of specific movement abnormalities before manifestation of gait impairment and quantification of disease progression in the prodromal phase.

**Methods:** 70 subjects participated in gait assessment, including 30 prodromal *SPAST* mutation carriers, 17 patients with mild-to-moderate manifest SPG4, and 23 healthy controls. Gait was assessed by an infrared-camera-based motion capture system to analyze features like range of motion and continuous angle trajectories. Those features were correlated with disease severity as assessed by the Spastic Paraplegia Rating Scale (SPRS) and neurofilament light chain (NfL) as a fluid biomarker indicating neurodegeneration.

**Results:** Compared to healthy controls, we found an altered gait pattern in prodromal mutation carriers during the swing phase in segmental angles of the lower leg (p<0.05) and foot (p<0.01), and in heel ground clearance (p<0.01). Furthermore, ranges of motion of segmental angles were reduced for foot (p<0.001) and lower leg (p<0.01). These changes occurred in prodromal mutation carriers without quantified leg spasticity in clinical examination. Gait features correlated with NfL levels and SPRS score.

**Conclusion:** Gait analysis can quantify changes in prodromal and mild-to-moderate manifest SPG4 patients. Thus, gait features constitute promising motor biomarkers characterizing the subclinical progression of spastic gait and might help to evaluate interventions in early disease stages.

## Introduction

Hereditary spastic paraplegias (HSP) are a clinically and genetically heterogeneous group of neurodegenerative disorders resulting in a length-dependent affection of the cortico-spinal tract (>80 HSP genes described^1^). Mutations in the *SPAST* gene result in spastic paraplegia type 4 (SPG4), comprising the most common cause of HSP^2^ and manifest as ‘pure’ HSP with hyperreflexia, leg spasticity, pyramidal weakness, and spastic gait resulting in increasing locomotor dysfunction^3, 4^.

As movement disturbance is critical in SPG4, gait features may serve as potential biomarkers to quantify gait abnormalities and disease progression in manifest disease and eventually already in the prodromal stage. Since the therapeutical potential of future interventions is likely most promising in the early stages of HSP^5^, it is crucial to identify and quantify the first changes already in the prodromal phase of genetically stratified cohorts.

Prodromal gait features have already been established in other neurodegenerative movement disorders like Parkinson’s disease^6^ and spinocerebellar ataxia^7, 8^. Thus far, gait analyses in HSP have been restricted to patients with clinically manifest gait abnormalities and patients with heterogeneous genetic background. In manifest HSP patients a reduced stride length and gait speed were observed as characteristic changes^9, 10^. Recent studies identified a decrease in range of motion (RoM) in the sagittal plane of joint and segmental angles^11, 12^. Serrao *et al*.^*11*^ clustered manifest HSP patients based on joint RoM values into severity-related groups^11^.

To establish movement biomarkers for characterizing the subclinical progression and natural history for possible therapeutic intervention effects in HSP, detailed knowledge about the gradual gait changes before manifestation of spatic gait is required. The prodromal period of disease provides a unique research opportunity to study such early gait alterations prospectively.

The *preSPG4 study* (NCT03206190) is an observational study which aims to characterize prodromal SPG4-related gait changes in prodromal and mild-to-moderate manifest SPG4 patients for comparison. We hypothesized that digital, objectively measured gait recording could quantify gradual gait changes already in the prodromal state of SPG4 when neither patients nor movement disorder specialists recognize spastic gait in clinical assessment.

## Materials and methods

### Participants

We included 17 SPG4 patients with manifest spastic gait disturbance from our spastic paraplegia outpatient clinic in Tübingen. All patients were able to walk without walking aids. In addition, 53 participants from the *preSPG4 study* (ClinicalTrials.gov Identifier: NCT03206190) were included. *preSPG4* is a single-center, rater-blinded, observational study in individuals at risk to develop SPG4. Descendants and siblings of SPG4 patients with a 50% risk to inherit the disease-causing *SPAST* mutation were included. The respective familial *SPAST* variant was analyzed to identify mutation carriers without disclosing the genetic result to the clinical examiners. Genetic analysis revealed 30 mutation carriers and 26 participants without a *SPAST* mutation. **SPG4 mutation carriers were categorized as prodromal** if HSP specialists rated their gait within the healthy spectrum (mean scores <2 points in the following scale: 0 ≙ normal gait, 1 ≙ could be within healthy spectrum, 2 ≙ could be within spastic spectrum, 3 ≙ definite spastic gait) in a blinded video assessment. The non-mutation carriers served as **healthy controls (HC)**. Clinical characterization and findings of the prodromal *SPAST* mutation carriers and healthy controls not carrying the *SPAST* mutation were reported in^13^. These mild-to-moderate **manifest** patients were used to define characteristics in digital, objectively measured gait abnormalities. Three healthy controls were excluded due to technical problems in the gait assessment resulting in 17 manifest SPG4 patients, 30 prodromal *SPAST* mutation carriers, and 23 healthy controls being analyzed. To assess the earliest stages of the disease, we defined a subgroup within the **prodromal SPG4** group with a cutoff in the SPRS^14^ score of <2, analogous to the range in SPRS of healthy controls (average + 2 SD)^14^. This prodromal **SPG4**_**SPRS<2**_ group included twelve individuals (nine without spasticity identified in the SPRS score). Characteristics of the four groups are shown in Table 1.

**Table 1:**
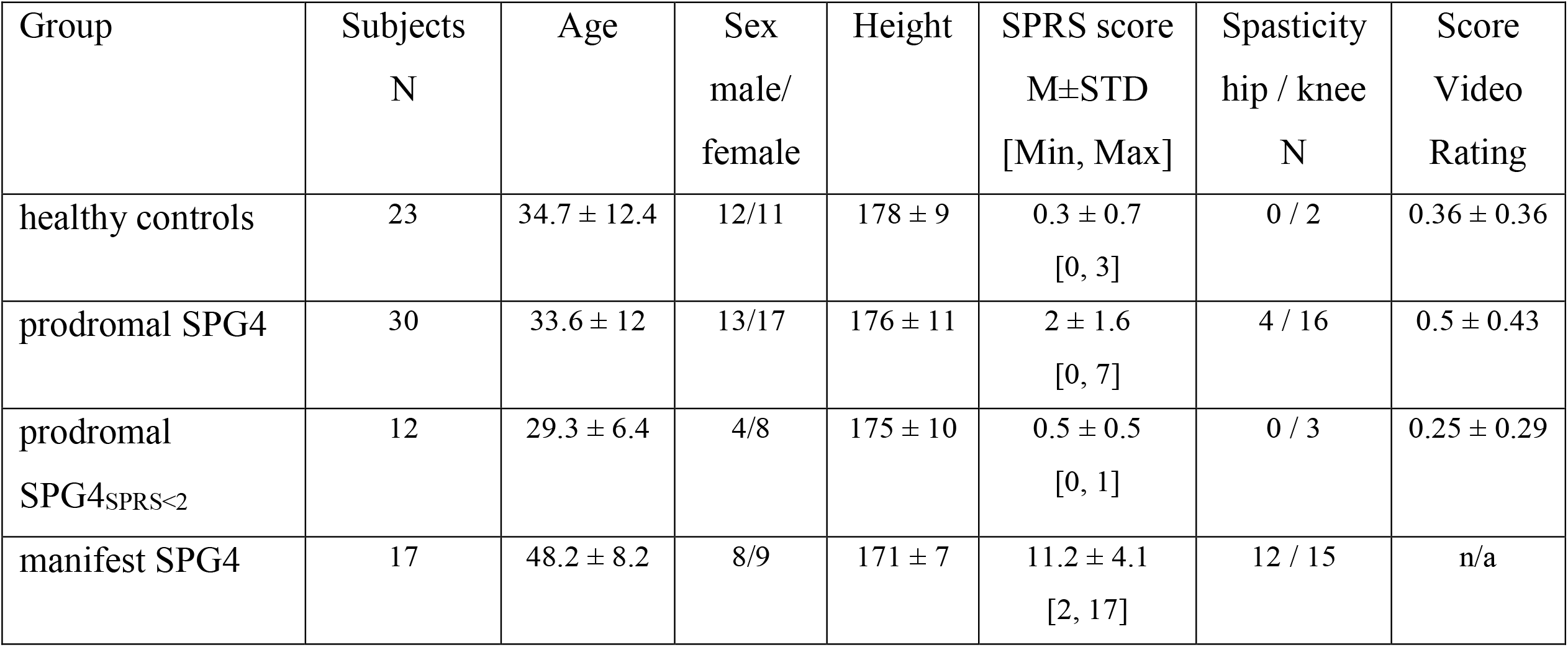
Demographic data of healthy controls, prodromal mutation carriers and manifest SPG4 patients. Shown are average values and standard deviation for mutation carriers and control groups. SPRS ≡ spastic paraplegia rating scale^14^; Spasticity ≡ Items 7 and 8 of SPRS, which represents spasticity of the hip adductors and knee extensors as measured by the modified Ashworth scale (mAS)^26^ with spasticity recognized as catch/release (mAS >0 points). N: Number. Reported mean video ratings with 0 ≙ normal gait, 1 ≙ could be within the healthy spectrum, 2 ≙ could be within the spastic spectrum, 3 ≙ definite spastic gait, n/a ≡ not applicable.

| Group | Subjects<br>N | Age | Sex<br>male/<br>female | Height | SPRS score<br>M±STD<br>[Min, Max] | Spasticity<br>hip / knee<br>N | Score<br>Video<br>Rating |
| --- | --- | --- | --- | --- | --- | --- | --- |
| healthy controls | 23 | 34.7 ± 12.4 | 12/11 | 178 ± 9 | 0.3 ± 0.7<br>[0, 3] | 0 / 2 | 0.36 ± 0.36 |
| prodromal SPG4 | 30 | 33.6 ± 12 | 13/17 | 176 ± 11 | 2 ± 1.6<br>[0, 7] | 4 / 16 | 0.5 ± 0.43 |
| prodromal<br>SPG4<sub>SPRS<2</sub> | 12 | 29.3 ± 6.4 | 4/8 | 175 ± 10 | 0.5 ± 0.5<br>[0, 1] | 0 / 3 | 0.25 ± 0.29 |
| manifest SPG4 | 17 | 48.2 ± 8.2 | 8/9 | 171 ± 7 | 11.2 ± 4.1<br>[2, 17] | 12 / 15 | n/a |

### Ethics approval

This study was carried out according to the Helsinki Declaration and approved by the Institutional Review Board of the University of Tübingen (reference numbers: 266/2017BO2) for the *preSPG4 study*. In addition, written informed consent was obtained from all study participants.

### Gait assessment and analysis

All participants underwent an instrumented gait analysis in a movement laboratory using an infrared-camera-based motion capture system (VICON FX with ten cameras). Subjects were instructed to walk normally (self-determined pace) with their own flat shoes for a 10m distance for several trials. Participants performed an average of 43 gait cycles. Three-dimensional movement trajectories were recorded with 41 reflecting markers, placed according to the Nexus Plug-in Gait model, at a sampling rate of 120 Hz.

Gait cycles were automatically extracted by detection of the heel strike events based on the heel marker positions’ vertical components for each foot and then verified manually using a stick figure animation to cross-check for different foot placements. Trials were smoothed with a Savitzky-Golay polynomial filter and resampled equidistantly with 101 data points per gait cycle by linear time interpolation. These procedures of gait analysis were used earlier in several studies on neurodegenerative movement disorders^15, 16^, including the quantification of pre-ataxic movement changes in spinocerebellar ataxia^8^.

As standard parameters, we computed gait parameters like speed, stride length, including the range of motion (RoM) of different segments (see Table 2), which have been identified as sensitive features in manifest HSP in earlier studies^9, 12, 17^. In addition, our analysis was focused on the segmental angle trajectories (see Figure 1 and Segmental Angles in the supplementary material) of the lower limb in the sagittal plane (flexion/extension) and leg marker position trajectories. Leg marker trajectories were normalized to a zero-ground level over the stance period to correct the individual characteristics of shoes. Each subject’s mean gait cycle trajectories were determined by averaging all gait cycles of the left and right leg. Furthermore, gait features were extracted from trajectories, e.g. maximum heel ground clearance or RoM of segmental angles.

**Table 2:** Gait features in healthy controls, prodromal mutation carriers, and manifest SPG4 patients. Analyzed gait measures for the four groups: healthy controls, prodromal SPG4 with SPRS <2, prodromal SPG4, and manifest SPG4 patients. Mean and standard deviation of features, p-values of Wilcoxon Ranksum test (*≡ p<0.05, **≡ p<0.0031 Bonferroni-corrected, ***≡ p<0.001) and Cliffs Delta as effect size (δ) is reported. n/a ≡ not applicable

| Gait measure | HC | prodromal<br>SPG4<sub>SPRS<2</sub> | prodromal<br>SPG4 | manifest<br>SPG4 | HC vs.<br>prodromal SPG4<sub>SPRS<2</sub> |  | HC vs.<br>prodromal SPG4 |  | HC vs.<br>manifest SPG4 |  | prodromal SPG4 vs.<br>manifest SPG4 |  |
| --- | --- | --- | --- | --- | --- | --- | --- | --- | --- | --- | --- | --- |
| | | | | | p | $\delta$ | p | $\delta$ | p | $\delta$ | p | $\delta$ |
| SPRS score | 0.3 ± 0.7 | 0.5 ± 0.52 | 2 ± 1.64 | 11.2 ± 4.13 | 0.13 | -0.26 | 0.000013*** | -0.67 | <0.00001*** | -1 | <0.00001*** | -0.94 |
| Serum NfL | 6.54 ± 2.36 | 7.46 ± 2.35 | 7.89 ± 3.59 | n/a | 0.27 | -0.23 | 0.18 | -0.22 | n/a | n/a | n/a | n/a |
| **Standard parameters** |  |  |  |  |  |  |  |  |  |  |  |  |
| Gait speed (m/s) | 1.36 ± 0.12 | 1.32 ± 0.11 | 1.28 ± 0.11 | 1.09 ± 0.21 | 0.66 | 0.09 | 0.037* | 0.34 | 0.000057*** | 0.75 | 0.0039* | 0.51 |
| Stride length (mm) | 1460 ± 90 | 1382 ± 127 | 1369 ± 113 | 1163 ± 190 | 0.085 | 0.36 | 0.005* | 0.45 | 0.00002*** | 0.8 | 0.00016*** | 0.67 |
| Gait cycle duration (s) | 1.08 ± 0.11 | 1.05 ± 0.06 | 1.07 ± 0.07 | 1.08 ± 0.14 | 0.24 | 0.25 | 0.69 | 0.07 | 0.87 | 0.03 | 0.97 | 0.01 |
| Step width (mm) | 88 ± 25.9 | 101 ± 28.9 | 95 ± 27.8 | 115 ± 40 | 0.29 | -0.22 | 0.55 | -0.1 | 0.033* | -0.4 | 0.09 | -0.31 |
| Circumduction(heel) % | 2.1 ± 2.38 | 4.2 ± 9.64 | 3.3 ± 6.61 | 2.2 ± 2.72 | 0.88 | -0.04 | 0.95 | -0.01 | 0.85 | 0.04 | 0.87 | 0.03 |
| Max ground clearance heel (cm) | 30.91 ± 2.71 | 28.43 ± 2.31 | 27.92 ± 2.35 | 24.77 ± 4.22 | 0.046* | 0.42 | 0.0017** | 0.51 | 0.00006*** | 0.75 | 0.01* | 0.46 |
| Max ground clearance toe (cm) | 9.98 ± 1.48 | 8.54 ± 1.26 | 8.99 ± 1.67 | 6.68 ± 2.61 | 0.01* | 0.54 | 0.027* | 0.36 | 0.00004*** | 0.77 | 0.0016** | 0.56 |
| **Segmental angles** |  |  |  |  |  |  |  |  |  |  |  |  |
| RoM Thigh (°) | 36.6 ± 2.55 | 35.1 ± 2.32 | 35.2 ± 3.08 | 34.6 ± 6.6 | 0.09 | 0.36 | 0.051 | 0.32 | 0.08 | 0.33 | 0.29 | 0.19 |
| RoM Lower Leg (°) | 75.4 ± 3.48 | 73.1 ± 2.79 | 72.5 ± 3.94 | 60.1 ± 12.79 | 0.042* | 0.43 | 0.0059* | 0.45 | 0.00002*** | 0.80 | 0.00019*** | 0.66 |
| RoM Foot (°) | 97.7 ± 4.42 | 91.8 ± 6.1 | 89.5 ± 7.92 | 70.8 ± 17.14 | 0.0087* | 0.55 | 0.00008*** | 0.63 | <0.00001*** | 0.92 | 0.00005*** | 0.72 |
| Min Thigh angle (°) | 78.6 ± 3.12 | 79.9 ± 2.33 | 79.7 ± 3.39 | 81 ± 4.23 | 0.21 | -0.27 | 0.26 | -0.18 | 0.04* | -0.38 | 0.18 | -0.24 |
| Max Thigh angle (°) | 115.2 ± 2.4 | 114.9 ± 2.9 | 114.9 ± 3.26 | 115.9 ± 3.99 | 0.88 | 0.04 | 0.78 | 0.05 | 0.37 | -0.17 | 0.35 | -0.17 |
| Min Lower Leg angle (°) | 34 ± 3.07 | 35.2 ± 2.01 | 34.9 ± 2.58 | 39.8 ± 7.84 | 0.23 | -0.25 | 0.21 | -0.20 | 0.0022** | -0.58 | 0.008* | -0.47 |
| Max Lower Leg angle (°) | 109.4 ± 2.83 | 108.3 ± 2.98 | 107.5 ± 3.28 | 99.9 ± 6.76 | 0.29 | 0.22 | 0.034* | 0.34 | <0.00001*** | 0.84 | 0.00005*** | 0.71 |
| Min Foot angle (°) | -70.5 ± 5.25 | -66.1 ± 6.15 | -63.4 ± 7.64 | -52.5 ± 12.06 | 0.042* | -0.43 | 0.00059*** | -0.56 | <0.00001*** | -0.89 | 0.0014** | -0.57 |
| Max Foot angle (°) | 27.3 ± 4.18 | 25.7 ± 3.75 | 26.2 ± 3.11 | 18.3 ± 7.94 | 0.32 | 0.21 | 0.23 | 0.19 | 0.0002*** | 0.7 | 0.0004*** | 0.63 |

**Figure 1:**
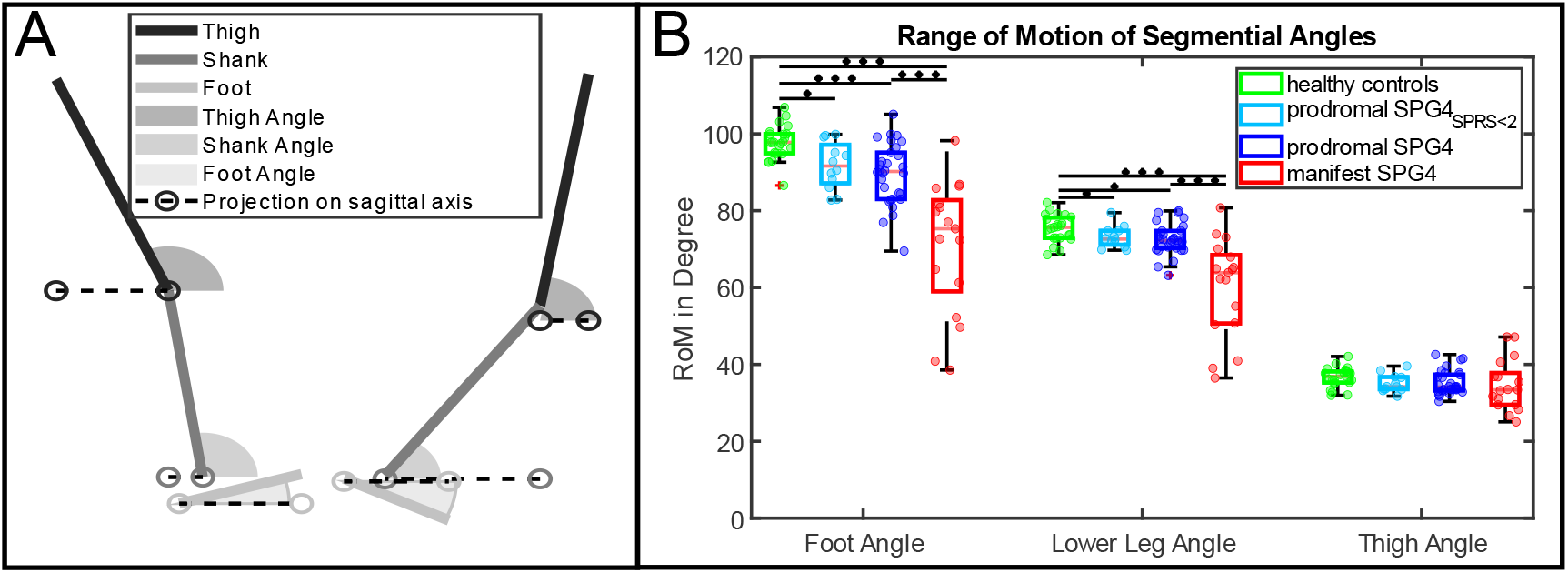
Range of motion of segmental angles in healthy controls, prodromal mutation carriers and manifest SPG4 patients. A) Calculation of segmental flexion angles in relation to the floor. Angles are always measured in reference to their projection on the sagittal axis, corresponding to the ground. Left side displays a positive foot angle, while the right side shows a negative foot angle. B) RoM of segmental angles of the four groups healthy controls, prodromal SPG4_SPRS<2_, prodromal SPG4, and manifest SPG4. Black bars indicate significance between groups with asterisks indicating: *≡ p<0.05, **≡ p<0.0031 Bonferroni-corrected, ***≡ p<0.001)

### Clinical Rating scales and fluid biomarkers

All participants were clinically examined by an HSP specialist (TWR), who rated disease severity according to the Spastic Paraplegia Rating Scale (SPRS)^14^.

Serum NfL levels, which were reported to be increased in manifest SPG4 patients^18^, were measured for all participants within the *preSPG4 study* (prodromal and healthy controls) as referred in^13, 19^.

### Statistics

The non-parametric Kruskal-Wallis-test was used to determine between-group differences in movement features. When the Kruskal-Wallis-test yielded a significant effect (p<0.05), posthoc analysis was performed using a Wilcoxon-Ranksum-test to compare groups. Effect sizes are reported by Cliff’s delta δ^20^ as small if δ<0.28, medium if 0.28<δ<0.43, and large if δ≥0.43, according to^21^. We report three significance levels: (i) uncorrected *: p<0.05, (ii) Bonferroni-corrected for multiple comparisons **: p<0.05/16=0.0031; n=16: number of analyzed features, (iii) ***: p<0.001. Linear regression was fitted to the stride length and each segmental angle’s RoM of prodromal SPG4 individuals to investigate the effect of stride length to segmental RoMs. We report coefficients of determination (R^2^) for comparison for the foot, lower leg, and thigh segmental RoM for prodromal SPG4 and healthy controls.

Wilcoxon-Ranksum-test was applied on each of the 101-time steps within one gait cycle for leg marker trajectories and segmental angles to estimate group differences.

We assessed four prominent gait cycle events: heel strike, heel-off, toe-off, and maximum heel ground clearance. Group differences for the segmental angles and marker trajectories were analyzed, and the most prominent event for each trajectory was used to correlate with the disease severity measured by SPRS score^14^. Identified gait features in the prodromal group were also correlated to NfL.

## Results

### Demographics

The manifest SPG4 group was significantly older than healthy controls and prodromal SPG4 patients (p=0.0013 and p<0.0001, respectively). There were no group differences between healthy controls and prodromal SPG4 in age and sex. For demographic details, see Table 1. The severity of the disease measured by the SPRS score differed significantly between all three groups (see Table 2 for more details).

### Gait changes in mild-to-moderate manifest SPG4 vs. healthy controls

Gait speed (p<0.001***, δ=0.75), stride length (p<0.001***, δ=0.8), and maximum heel and toe ground clearance (p<0.001***, δ=0.75 and p<0.001***, δ=0.77, respectively) were significantly reduced in mild-to-moderate manifest SPG4 compared to healthy controls. RoM of the foot (p<0.001***, δ=0.92) and lower leg (p<0.001***, δ=0.8) were also reduced significantly (Figure 1 B). Further, maximum and minimum angles in the foot (δ=0.7, δ=-0.89) and lower leg (δ=0.84, δ=-0.58) were reduced/increased significantly (p<0.001***), see Table 2 and Figure 2 for more details.

**Figure 2:**
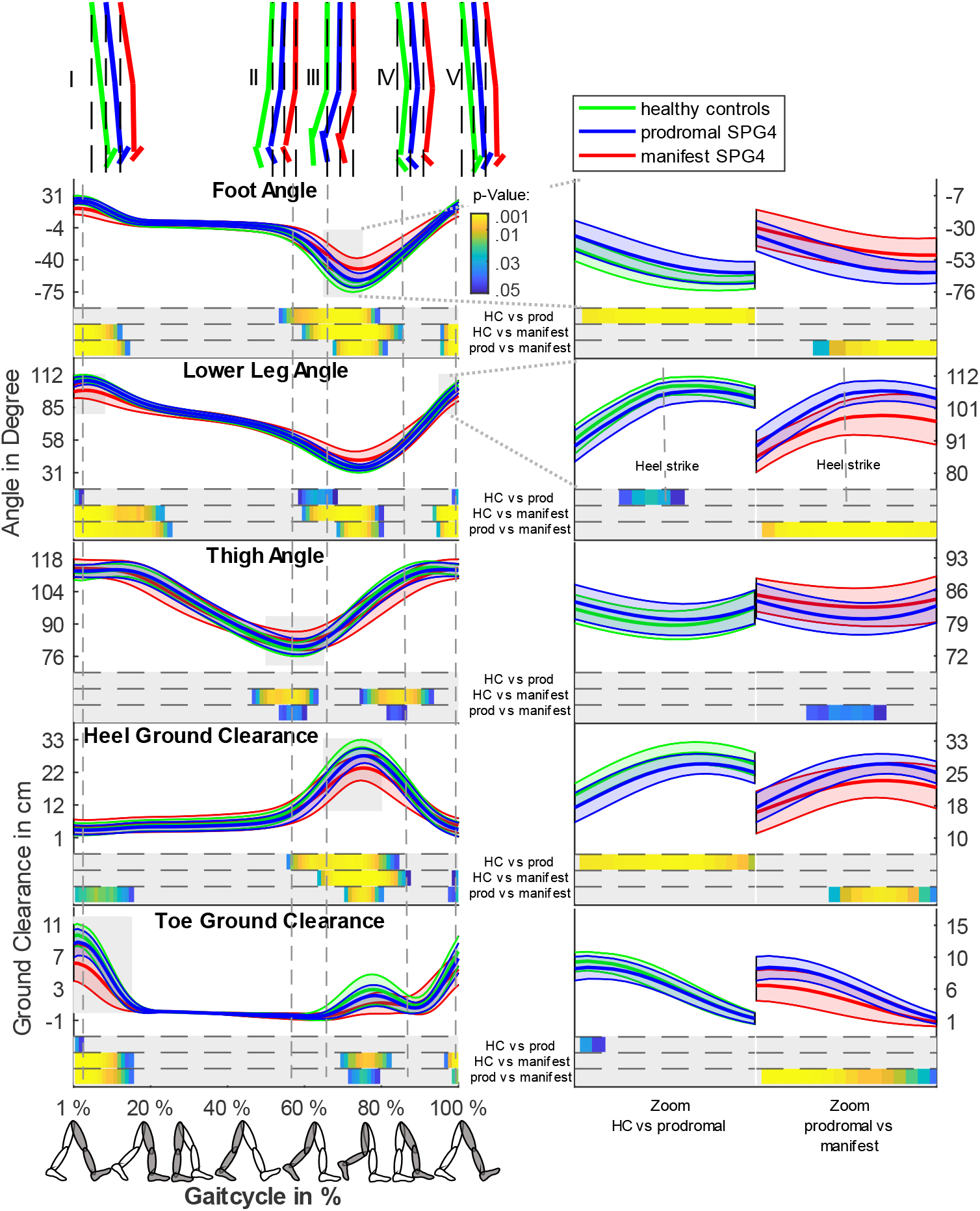
Gait trajectories and segmental angles during a gait cycle of HC, prodromal SPAST mutation carriers and manifest SPG4 patients. Individual trajectories and segmental angles during a gait cycle (from heel strike to heel strike) for groups healthy controls (green), prodromal SPG4 (blue) and manifest SPG4 (red). On the left side of the figure, the whole gait cycle is displayed for the three segmental angles and the heel and ankle ground clearance. In the upper part, the thigh, lower leg, and foot segments are plotted based on the mean angles to the given time point (dashed line). Wilcoxon Ranksum was applied to each percent step of the gait cycle to compare group differences and significant results (p < 0.05) are displayed color-coded below each feature. Group-wise comparison is displayed as healthy controls vs prodromal SPG4, healthy controls vs manifest SPG4 and prodromal SPG4 vs manifest SPG4. On the right side, outtakes of the gait cycle around the most characteristic time points are displayed separately for healthy controls vs prodromal SPG4 and prodromal SPG4 vs manifest SPG4. The corresponding gait period is shaded grey on the left side.

### Gait changes in prodromal *SPAST* mutation carriers vs. healthy controls

Prodromal mutation carriers compared to healthy controls revealed reduced maximum ground clearance of heel (p=0.0017**, δ=0.51) and toe (p=0.026*, δ=0.36), stride length (p=0.005*, δ=0.45), and gait speed (p=0.037*, δ =0.34). RoMs of the foot (p<.001***, δ=0.63) and lower leg (p=0.0059**, δ=0.45) segmental angles were significantly reduced, as shown in Figure 1 B. Coefficients of determination for the foot (R^2^_prodromal SPG4_ = 0.21; R^2^_HC_ = 0.03), the lower leg (R^2^_prodromal SPG4_ = 0.21; R^2^ _HC_ = 0.03), and the thigh (R^2^_prodromal SPG4_ = 0.11; R^2^ _HC_ = 0.18) segments, indicate that the decrease of the segmental angles RoMs were not predominantly driven by stride length.

The RoM effect size of the lower leg angle (δ=0.63) was comparable to the effect size of the SPRS score (δ=0.67). Further results and p-values are shown in Table 2.

The in-depth analysis of differences within a gait cycle revealed specific movement changes of prodromal SPG4: Differences occurred mainly in the swing phase and around the heel strike event. During the swing phase, the magnitudes of foot and lower leg angle and heel ground clearance were reduced (Figure 2 (III-IV)). At the heel strike event, the lower leg angle and toe ground clearance were significantly reduced in prodromal mutation carriers (Figure 2 (I & V)).

### First gait signs in prodromal SPG4_SPRS<2_

Remarkably, differences were identified even for the prodromal subgroup **SPG4**_**SPRS<2**_ compared to healthy controls. RoM in segmental angle of the foot (p=0.0087*, δ=0.55) and lower leg (p=0.042*, δ=0.43) were significantly reduced as well as maximum heel (p=0.046*, δ=0.42) and toe (p=0.01*, δ=0.54) ground clearance. In the prodromal **SPG4**_**SPRS<2**_ subgroup, effect sizes in all significant gait features were substantially larger (up to 210%) than for the SPRS score (δ=-0.26). In contrast to the whole prodromal SPG4 group and manifest SPG4 patients, stride length did not differ between prodromal **SPG4**_**SPRS<2**_ and healthy controls. Differences at heel strike, heel-off, toe-off, and maximum ground clearance of the heel are shown in Figure 3 depicted for all four groups.

**Figure 3:**
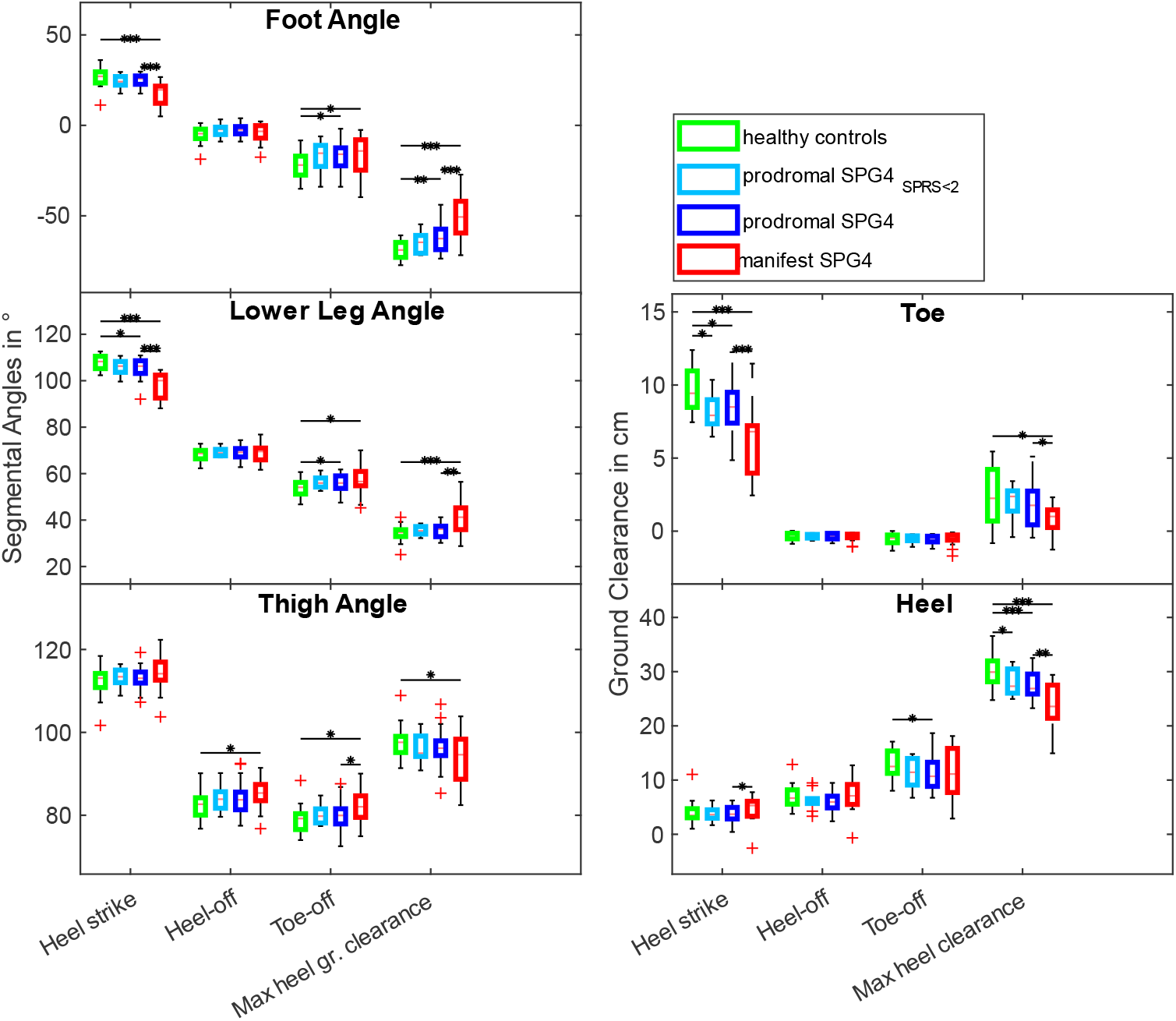
Gait features within the gait cycle of HC, prodromal *SPAST* mutation carriers and patients with manifest SPG4. Boxplots of Foot Angle, Lower Leg Angle, Thigh Angle, Toe Marker, and Heel Marker at the heel strike, heel-off, toe-off, and at maximum heel ground clearance. Displayed are the four groups: healthy controls, prodromal SPG4_SPRS<2_, prodromal SPG4, and manifest SPG4. Black bars indicate significance between groups with asterisks indicating: *≡ p<0.05, **≡ p<0.0031 Bonferroni-corrected, ***≡ p<0.001)

### Correlations of gait features with severity of disease

We chose the three segmental angles (foot, lower leg, thigh) and ground clearance (toe and heel) at prominent events within the gait cycle (heel strike, heel-off, toe-off, and max heel ground clearance). Correlations between these gait features and disease severity (SPRS score) in patients with manifest SPG4, prodromal mutation carriers, and healthy controls are shown in Supplementary Figure 1. Within the whole range of disease severity (SPRS range 0-17), all chosen gait features correlated significantly with the SPRS score (e.g., foot angle at maximum heel ground clearance: p<0.001***).

In patients with manifest SPG4, the foot (r=-0.57, p=0.016*), lower leg (r=-0.55, p=0.023*), and thigh RoM (r=-0.6, p=0.012*), the foot angle at maximum heel ground clearance (r=0.63, p=0.006*) the lower leg angle at heel strike (r=-0.49, p=0.04*), and thigh angle at toe-off (r=0.59, p=0.012*) correlated with the SPRS score.

In the prodromal group, gait characteristics were shifted with disease progression gradually towards the abnormalities seen in mild-to-moderate manifest SPG4 patients, as illustrated in Figure 2 by the blue line (prodromal mutation carriers) passing between those of healthy controls (green line) and manifest SPG4 patients (red line). The foot angle at maximum heel ground clearance (r=0.58, p<0.001***) and the foot RoM (r=-0.49, p=0.005*) of prodromal SPG4 correlated significantly with the SPRS score.

Moreover, for prodromal mutation carriers the foot angle at maximum heel ground clearance (r=0.48, p=0.0075*) and the foot RoM (r=-0.46, p=0.01*) correlated significantly with NfL levels in serum. In contrast, healthy controls showed no correlation (foot angle: r=0.18, p=0.58; foot RoM: r=0.12, r=0.52).

## Discussion

This study analyzed gait in the prodromal and mild-to-moderate manifest stage of SPG4, the most common subtype of hereditary spastic paraplegia. We found characteristic gait abnormalities in patients with mild-to-moderate manifest SPG4 and proved significant changes in gait parameters already in the prodromal stage of disease before movement specialists recognized gait abnormalities and before clinical assessment (SPRS) shows discriminative results from healthy controls (SPG4_SPRS<2_ subgroup). The quantified changes showed large effect sizes, which are essential for motor biomarkers in rare diseases like SPG4 to achieve high statistical power despite small cohorts in upcoming clinical trials.

### Gait characteristics in mild-to-moderate manifest SPG4

Characteristic gait changes in mild-to-moderate manifest SPG4 patients (SPRS 2-17) were a significant reduction of foot and lower leg RoM resulting from both increased minimum and decreased maximum of the segmental angles. The reduction in RoM of segmental angles^9, 11, 12^, step height (heel ground clearance), gait speed, and stride length are in line with previous reports^10, 22^. The observed reduced RoMs in lower limb segments are supposed to be caused by an interplay of lower limb spasticity and pyramidal weakness, two primary HSP symptoms. We found reduced foot ground clearance during the swing phase of a gait cycle, which supports Kerstens *et al*.^23^ qualitative findings that stumbling is a crucial feature in manifest HSP locomotion and gait measures correlate with fear of falling^10^.

Besides identifying gait characteristics that distinguished mild-to-moderate manifest patients from healthy controls, we searched for gait measures that revealed sensitivity to disease severity change. In line with disease severity correlations in the study of Martino *et al*.^12^, the SPRS score correlated with the RoMs of the foot and lower leg angle in our study. The reduced levels of joint RoM were previously used by Serrao *et al*.^11^, to cluster three different groups of manifest HSP patients. Our observation of reduced ROMs in foot and lower leg correspond to the intermediate severe group in^11^ (SPRS score: 16.07 ± 6.48).

Regarding specific events in the gait cycle, we found correlations of the the SPRS score with foot angle at maximum heel ground clearance (r=0.63, p=0.006*), the lower leg angle at the heel strike event (r=-0.49, p=0.04*), and the thigh angle at the toe-off event (r=0.59, p=0.012*). These events highlight features immediately after fast joint flexion/extension periods during the gait cycle (see Figure 2, instant **III-IV** for foot angle, instant **I** for lower leg angle, and instant **III** for thigh angle). These observations would fit to the hypothesis, that fast muscle contraction during swing (e.g., gastrocnemius and soleus) leads to spasticity and thus to a reduced mobility in the foot angle. The reduced segmental angles correlated with the SPRS score; this may be explained by the velocity-dependent stiffness of the joints HSP^24^. The reduction of the segmental angles and RoMs in manifest patients are supposed to be driven by muscle paresis, especially of the dorsiflexor and hip flexor muscles, combined with severe stiffness of plantar flexor and knee extensor muscles^24^, which are symptoms that are also assessed by the SPRS^14^. For all manifest patients, spasticity and/or pyramidal weakness was documented in at least one joint in the SPRS. These results indicated that the progression of mild-to-moderate manifest SPG4 patients who can still walk freely (without walking aids) could be quantified due to spasticity-dependent outcomes in the gait cycle.

### Gait characteristics in prodromal SPG4

In prodromal SPG4, the range of motion of the foot and lower leg segments (Figure 1), stride length, gait speed, and maximum heel and toe ground clearance were reduced. These changes represent the gradual appearance of those seen in manifest SPG4 patients. Similar to the mild-to-moderate manifest SPG4 patients, the thigh RoM was not reduced for prodromal SPG4; however, less spasticity in the hip joint was apparent in the spasticity assessment of the SPRS in our study (13% prodromal SPG4 and 72% manifest SPG4 with spasticity in the hip joint). These differences were visible during the gait cycle at the toe-off event and during the swing phase (Figure 2, instances II and III), showing the importance of identifying relevant time points within the gait cycle. In comparison to hip spasticity, knee spasticity was already identified in 53% of prodromal SPG4 (see Table 1), supporting that distal muscles are more affected^24^, due to length-dependent affection of the cortico-spinal tract.

Importantly, significant gait changes could already be quantified for the prodromal subgroup SPG4_SPRS<2_, affecting the foot and lower leg RoM, and the minimum foot plantarflexion. These changes in the segmental angles were already reflected in the gradually reduced foot ground clearance during the swing phase. The changes in this subgroup (i.e., foot and lower leg RoM with effect sizes δ=0.55 and δ=0.43, respectively) highlight the relevance of instrumented gait analyses in the very early phase of SPG4 since it quantifies the earliest changes that were neither visible for movement disorder specialists nor identified by the clinical SPRS score.

### Gait features as a biomarker in the prodromal SPG4

In the prodromal disease stage, quantifiable biomarkers are essential as possible study endpoints for therapeutic endeavors. Our results suggest that gait features can fulfill these requirements by revealing gradual progression during the prodromal phase of SPG4, with the foot RoM and foot segmental angle at the maximum heel ground clearance and minimum plantarflexion correlating with first disease symptoms (measured by the SPRS). The hypothesis that the identified gait changes are indeed an expression of incipient changes in SPG4 with specific neural degeneration is strengthened by the correlation of gait parameters with NfL values in the prodromal phase. Neurofilament light chain (NfL) is a fluid biomarker indicating axonal degeneration processes^18, 25^ and is increased in prodromal SPG4 and manifest SPG4 patients^13^. Two of our most sensitive gait measures in the prodromal phase showed significant correlations to NfL levels in serum (foot angle at maximum heel ground clearance: r=0.48, p=0.0075* and the foot RoM: r=-0.46, p=0.01*) for the prodromal group. Thus, these gait measures present a promising performance measure associated with neural degeneration in the prodromal and mild-to-moderate manifest phase of SPG4, including subjects without visible spastic gait.

## Conclusion

This study was able to quantify subclinical gait impairment in *SPAST* mutation carriers even without any clinical sign of pyramidal affection. Objectively measured gait features indicate disease progression over the prodromal and mild-to-moderate manifest phase and revealed larger effect sizes than clinical scores for the mildest affected subgroup, potentially reducing sample size in future interventional trials. Thus, quantitative movement features represent promising candidates for motor biomarkers in future therapeutic interventions in homogenous cohorts of prodromal SPG4. We will further investigate the changes in gait during disease progression in longitudinal assessments to define the sensitivity to change of movement markers in the early stages of SPG4.

## Data Availability

The data sets for this manuscript are not publicly available because raw data regarding human subjects (e.g., genetic raw data, personal data) are not shared freely to protect the privacy of the human subjects involved in this study; no consent for open sharing has been obtained. Requests to access an anonymous data set should be directed to Dr. Winfried Ilg and Dr. Tim W. Rattay.

## Abbreviations

HSP: Hereditary spastic paraplegia
RoM: Range of motion
SPG4: Spastic paraplegia gene type 4
SPRS: Spastic Paraplegia Rating Scale
NfL: Neurofilament light chain

## Acknowledgments

LS is a member of the European Reference Network for Rare Neurological Diseases - Project ID No 739510. We thank Holger Hengel (HH) for the video gait ratings of the preSPG4 probands and Katrin Dillman-Jehn for the help in coordinating communication with suitable families and probands.

## Author Roles

1. Research project: A. Conception, B. Organization, C. Execution
2. Statistical Analysis: A. Design, B. Execution, C. Review and Critique
3. Manuscript Preparation: A. Writing of the first draft, B. Review and Critique

C.L.: 1B, 1C, 2A, 2B, 3A

W.I.: 1A, 1B, 2A, 2C, 3B

M.S.: 1B, 1C, 3B

M.V.: 1B, 3B D.F.B.H.: 2C, 3B

M.G.: 1B, 2C, 3B

R.S.: 1B, 2C, 3B

L.S.: 1A, 2C, 3B

T.W.R.: 1A, 1B, 2C, 3B

## Supplementary Material

**Supplementary Figure 1:**
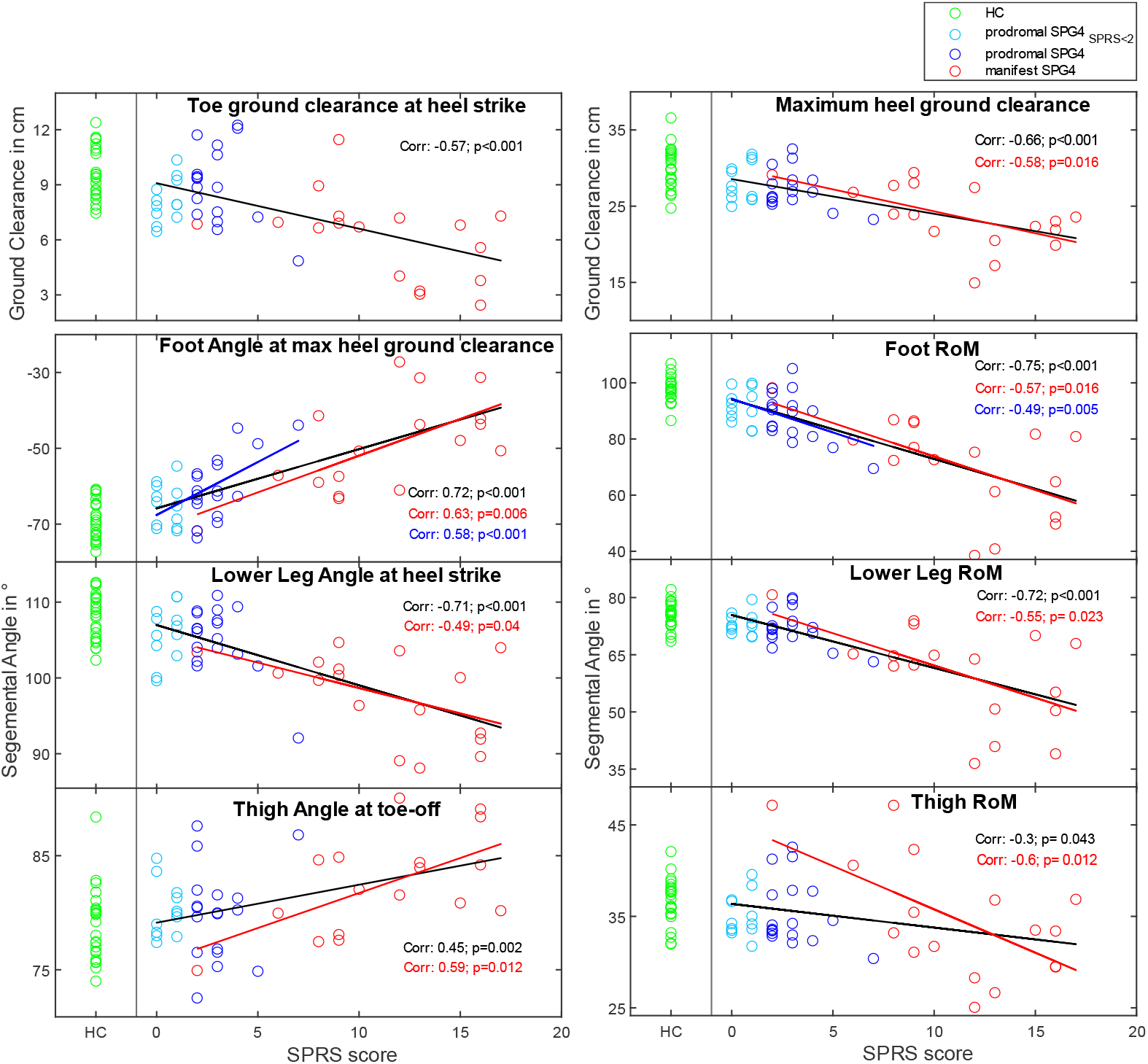
Correlations of gait features at events within the gait cycle against SPRS score. Plots of gait features at events within the gait cycle against SPRS score showing the correlations of gait features and disease severity. The top row shows toe and heel ground clearance. Left column bottom three plots show segmental angles at prominent events, right column bottom three plots showing segmental angles range of motions. The black line indicates linear fit for all SPG4 groups (prodromal and manifest) together with significant correlations. The red and blue lines show linear fit for manifest SPG4 only and prodromal only with significant correlation. Healthy controls were plotted for comparison.

## Supplementary Explanation 1: Calculation of segmental angles

Calculating the segmental angles in relation to the ground reflects the summative changes in posture. Therefore, the foot segmental angle shows the sum of the orientation of the upper body and the hip, knee, and ankle joint angles. Angles of limb segments were previously used by Martino *et al*.^*12*^ in HSP patients and introduced by Borghese *et al*.^27^.

The segmental angles were determined for the thigh, lower leg, and foot and calculated as the angle between the segment on the longitudinal and sagittal axes and its projection on a sagittal axis, corresponding to the ground (see Figure 1 A). Calculation of the flexion angles of the segments was based on the sagittal and longitudinal marker positions.

The thigh segment’s upper end was calculated as the average between the posterior superior iliac and the anterior superior iliac. For the lower end, the knee marker was used. The lower leg segment was defined by the knee and ankle marker positions, corresponding to the upper and lower end of the lower leg, respectively. The foot segment was determined by the heel and toe marker positions. The foot angle can be positive (heel below the toe on the longitudinal axis) and negative (heel above toe on the longitudinal axis), as shown in Figure 1.

## References

1. Bis-Brewer DM, Zuchner S. Perspectives on the Genomics of HSP Beyond Mendelian Inheritance. Front Neurol 2018;9:958.

2. Hazan J, Fonknechten N, Mavel D, et al. Spastin, a new AAA protein, is altered in the most frequent form of autosomal dominant spastic paraplegia. Nat Genet 1999;23(3):296–303.

3. Harding AE. Classification of the hereditary ataxias and paraplegias. Lancet 1983;1(8334):1151–1155.

4. Fink JK. Hereditary spastic paraplegia. Current neurology and neuroscience reports 2006;6(1):65–76.

5. Lallemant-Dudek P, Darios F, Durr A. Recent advances in understanding hereditary spastic paraplegias and emerging therapies. Fac Rev 2021;10:27.

6. Mirelman A, Gurevich T, Giladi N, Bar-Shira A, Orr-Urtreger A, Hausdorff JM. Gait alterations in healthy carriers of the LRRK2 G2019S mutation. Annals of neurology 2011;69(1):193–197.

7. Rochester L, Galna B, Lord S, Mhiripiri D, Eglon G, Chinnery PF. Gait impairment precedes clinical symptoms in spinocerebellar ataxia type 6. Movement Disorders 2014;29(2):252–255.

8. Ilg W, Fleszar Z, Schatton C, et al. Individual changes in preclinical spinocerebellar ataxia identified via increased motor complexity. Mov Disord 2016;31(12):1891–1900.

9. Piccinini L, Cimolin V, D’Angelo MG, Turconi AC, Crivellini M, Galli M. 3D gait analysis in patients with hereditary spastic paraparesis and spastic diplegia: a kinematic, kinetic and EMG comparison. Eur J Paediatr Neurol 2011;15(2):138–145.

10. Gaßner H, List J, Martindale CF, et al. Functional gait measures correlate to fear of falling, and quality of life in patients with Hereditary Spastic Paraplegia: A cross-sectional study. Clinical Neurology and Neurosurgery 2021;209:106888.

11. Serrao M, Rinaldi M, Ranavolo A, et al. Gait Patterns in Patients with Hereditary Spastic Paraparesis. PLOS ONE 2016;11(10):e0164623.

12. Martino G, Ivanenko Y, Serrao M, et al. Locomotor coordination in patients with Hereditary Spastic Paraplegia. Journal of Electromyography and Kinesiology 2019;45:61–69.

13. Rattay TW, Völker M, Rautenberg M, et al. The prodromal phase of Hereditary Spastic Paraplegia Type 4 – the preSPG4 cohort study. Brain 2022;accepted.

14. Schüle R, Holland-Letz T, Klimpe S, et al. The Spastic Paraplegia Rating Scale (SPRS): a reliable and valid measure of disease severity. Neurology 2006;67(3):430–434.

15. Ilg W, Christensen A, Mueller OM, Goericke SL, Giese MA, Timmann D. Effects of cerebellar lesions on working memory interacting with motor tasks of different complexities. Journal of Neurophysiology 2013;110(10):2337–2349.

16. Ilg W, Schatton C, Schicks J, Giese MA, Schöls L, Synofzik M. Video game–based coordinative training improves ataxia in children with degenerative ataxia. Neurology 2012;79(20):2056–2060.

17. Martino G, Ivanenko Y, Serrao M, et al. Differential changes in the spinal segmental locomotor output in Hereditary Spastic Paraplegia. Clinical Neurophysiology 2018;129(3):516–525.

18. Kessler C, Serna-Higuita LM, Wilke C, et al. Characteristics of serum neurofilament light chain as a biomarker in hereditary spastic paraplegia type 4. Annals of Clinical and Translational Neurology 2022;9(3):326–338.

19. Wilke C, Rattay TW, Hengel H, et al. Serum neurofilament light chain is increased in hereditary spastic paraplegias. Annals of Clinical and Translational Neurology 2018;5(7):876–882.

20. Cliff N. Dominance statistics: Ordinal analyses to answer ordinal questions. Psychological Bulletin 1993;114(3):494–509.

21. Vargha A, Delaney HD. A Critique and Improvement of the CL Common Language Effect Size Statistics of McGraw and Wong. Journal of Educational and Behavioral Statistics 2000;25(2):101–132.

22. Klebe S, Stolze H, Kopper F, et al. Gait analysis of sporadic and hereditary spastic paraplegia. Journal of Neurology 2004;251(5):571–578.

23. Kerstens HCJW, Satink T, Nijkrake MJ, et al. Stumbling, struggling, and shame due to spasticity: a qualitative study of adult persons with hereditary spastic paraplegia. Disability and Rehabilitation 2020;42(26):3744–3751.

24. Marsden J, Ramdharry G, Stevenson V, Thompson A. Muscle paresis and passive stiffness: Key determinants in limiting function in Hereditary and Sporadic Spastic Paraparesis. Gait & Posture 2012;35(2):266–271.

25. Kessler C, Serna-Higuita LM, Rattay TW, et al. Neurofilament light chain is a cerebrospinal fluid biomarker in hereditary spastic paraplegia. Annals of Clinical and Translational Neurology 2021;8(5):1122–1131.

26. Bohannon RW, Smith MB. Interrater Reliability of a Modified Ashworth Scale of Muscle Spasticity. Physical Therapy 1987;67(2):206–207.

27. Borghese NA, Bianchi L, Lacquaniti F. Kinematic determinants of human locomotion. The Journal of Physiology 1996;494(3):863–879.

